# Discrepancies in automated, electronic medical record-based CHA2DS2-VASc scores and clinician assessment for atrial fibrillation patients

**DOI:** 10.1101/2020.12.23.20248775

**Authors:** Aubrey E. Jones, Zameer Abedin, Olesya Ilkun, Rebeka Mukherjee, Mingyuan Zhang, Michael White, Benjamin A. Steinberg, Rashmee U. Shah

## Abstract

**Background:** Clinical decision support tools for atrial fibrillation (AF) should include CHA_2_DS_2_- VASc scores to guide oral anticoagulant (OAC) treatment.

**Objective:** We compared automated, electronic medical record (EMR) generated CHA_2_DS_2_- VASc scores to clinician-documented scores, and report the resulting proportions of patients in the OAC treatment group.

**Methods:** Patients were included if they had both a clinician documented and EMR-generated CHA_2_DS_2_-VASc score on the same day. EMR scores were based on billing codes, left ventricular ejection fraction from echocardiograms, and demographics; documented scores were identified using natural language processing. Patients were deemed “re-classified” if the EMR score was ≥2 but the documented score was <2, and vice versa. For the overall cohort and subgroups (sex and age group), we compared mean scores using paired t-tests and re-classification rates using chi-squared tests.

**Results:** Among 5,767 patients, the mean scores were higher using EMR compared to documented scores (4.05 [SD 2.1] versus 3.13 [SD 1.8]; p<0.01) for the full cohort, and all subgroups (p<0.01 for all comparisons). If EMR scores were used to determine OAC treatment instead of documented scores, 8.3% (n=479, p<0.01) of patients would be re-classified, with 7.2% moving into and 1.1% moving out of the treatment group. Among 2,322 women, 4.7% (n=109, p<0.01) would be re-classified, with 4.1% into and 0.7% out of the treatment group. Among 3,445 men, 10.7% (n=370, p<0.01) would be re-classified, with 9.2% into and 1.5% out of the treatment group. Among 2,060 patients <65 years old, 18.1% (n=372, p<0.01) would be re-classified, with 15.8% into and 2.3% out of the treatment group. Among 1,877 patients 65-74 years old, 5.4% (n=101, p<0.01) would be re-classified, with 4.4% into and 1.0% out of the treatment group. Among 1,830 patients ≥75 years old, <1% would move into to the treatment group and none would move out of the treatment group.

**Conclusions:** EMR-based CHA_2_DS_2_-VASc scores were, on average, almost a full point higher than the clinician-documented scores. Using EMR scores in lieu of documented scores would result in a significant proportion of patients moving into the treatment group, with the highest re-classifications rates in men and patients <65 years old.

## Introduction

Atrial fibrillation (AF) is the most common heart arrhythmia and is associated with an increased risk of stroke and other thromboembolic events. While oral anticoagulation (OAC) significantly decreases this risk, treatment comes with an increased risk of bleeding. Therefore, risk stratification using the CHA_2_DS_2_-VASc score is used to ensure that only patients with a high risk of stroke—enough to warrant the bleeding risk— receive anticoagulation.^1^ The CHA_2_DS_2_- VASc score ranges from 0 to 9, and includes age, sex, and five comorbid conditions: heart failure, hypertension, diabetes mellitus, vascular disease, and prior thromboembolic event. Calculating the CHA_2_DS_2_-VASc score, however, can be challenging for clinicians due to time constraints; manually scoring patients one by one makes population level characterization impractical. A more efficient method to generate CHA_2_DS_2_-VASc scores at the point of care and across populations is needed.

Previous studies have shown that 50% to 60% of AF patients at an increased risk of stroke (CHA_2_DS_2_-VASc score ≥2) are not treated with OAC.^2,3^ Moreover, inappropriate treatment with OAC exposes patients to unnecessary, increased risk of bleeding. By accurately determining the population at risk, more meaningful and targeted interventions can be made in order to optimize clinical outcomes for these patients. Electronic medical records (EMRs) include a substantial amount of data that could be used to automatically calculate CHA_2_DS_2_- VASc scores across large patient populations, and help improve OAC treatment rates. The goal of these analyses was to: (1) assess agreement between EMR-based (EMR) and clinician-documented (documented) scores; (2) determine if EMR scores were more likely to be higher or lower than documented CHA_2_DS_2_-VASc scores; and (3) assess the potential impact on OAC treatment rates by using EMR versus documented scores.

## Methods

This was a retrospective study comparing two different methods of calculating the CHA_2_DS_2_-VASc score. The candidate population included AF patients at University of Utah Health with at least one EMR, automated (EMR score) and one clinician, documented (documented score) CHA_2_DS_2_-VASc score from the same day. If patients had more than one score documented on the same day, the higher of the two scores was used. If patients had more than one set of scores from different days, we included the one closest in time to the first AF diagnosis.

Our health system previously developed a process to automatically generate CHA_2_DS_2_- VASc scores for AF patients using demographics, International Classification of Diseases (ICD) diagnosis billing codes, and additional echocardiographic. ^4,5^ ICD Version 9 and 10 codes from the patients’ entire record within the system were mapped to each of the conditions in the CHA_2_DS_2_-VASc score. In addition to ICD codes, patients with a left ventricular ejection fraction ≤40% (by echocardiogram) within the 365-day period prior to the date of the patient visit under consideration were considered to have heart failure. Comorbidity component scores were tabulated where the earliest date of comorbidity diagnosis was less than or equal to the date of the patient visit under consideration. Age and gender component scores were added to the corresponding comorbidity component scores to arrive at the final EMR score for each patient visit.

For comparison, we used natural language processing (NLP) to extract clinician documented CHA_2_DS_2_-VASc scores from the clinical notes (e.g. “CHA_2_DS_2_-VASc score is 4, continue warfarin”). We did not target or include CHADS_2_ scores, an older method of stroke risk assessment. We identified a list of target terms that clinicians use to reference CHA_2_DS_2_-VASc scores and the computer algorithm parsed each phrase within a note for every patient, using regular expression matching to find occurrences of target terms. For each target term, the algorithm looked for a number in the text that occurred within 15 characters after the identified target term. This number was captured as the CHA_2_DS_2_-VASc score documented by the clinician. If an additional CHA_2_DS_2_-VASc target term was identified within 15 characters, numbers were extracted only up to the next target term. To assess the accuracy of the NLP system, we randomly selected 33 phrases that included references to a CHA_2_DS_2_-VASc score. We manually reviewed these phrases and found that the algorithm was incorrect in 2 of 33 cases, for an accuracy of 94%.

We compared EMR and documented scores using a paired t-test. We also calculated the number of patients who would move into and out of the guideline-directed OAC treatment group (“re-classification”) if using automated scores, CHA_2_DS_2_-VASc score of ≥2, which was the prevailing cut-off over the study period.^6^ Chi^2^ tests of significance were used for all categorical variables. Additionally, subgroup analyses were performed evaluating differences in CHA_2_DS_2_- VASc score between the two methods, stratified by sex and age group (≤64, 65-74, ≥75 years old). Analyses were performed using Stata and the Python programming language. This retrospective, quality improvement study was approved by the Institutional Review Board at the University of Utah.

## Results

We identified 5,767 adult AF patients seen between 2010 and 2017 with an EMR and a documented CHA_2_DS_2_-VASc score on the same day. The mean age of the population was 67.7 years old (SD 12.8), 40.3% were female, and 90.2% were White (Table 1). Overall, the agreement between the exact EMR and documented scores was 42.3%. However, when the scores were classified into two treatment groups, 0-1 or ≥2, the scores agreed 91% of the time. The mean EMR CHA_2_DS_2_-VASc score was about one point higher than the mean documented score (4.05 [SD 2.1] versus 3.13 [SD 1.8]; p<0.01; Figure 1).

**Table 1:** Basic characteristics of 5,767 atrial fibrillation patients with a clinician documented and EMR generated CHA_2_DS_2_-VASc score on the same day.

| Characteristic<sup>1</sup> | Prevalence, n (%) |
| --- | --- |
| Female | 2,322 (40.3) |
| Age mean, (SD) | 67.7 (12.8) |
| Age ≤64 years old | 2,060 (35.7) |
| Age 65-74 years old | 1,877 (32.5) |
| Age ≥75 years old | 1,830 (31.7) |
| Race |  |
| White | 5,199 (90.2) |
| Black | 44 (0.7) |
| Asian | 63 (1.1) |
| Other/missing | 461 (8.0) |
| Comorbidities<sup>2</sup> |  |
| Acute myocardial infarction | 502 (8.7) |
| Coronary artery disease | 1,922 (33.3) |
| Congestive heart failure | 1,499 (26) |
| Diabetes mellitus | 1,723 (29.9) |
| Cerebrovascular disease | 952 (16.5) |
| Hypertension | 3,673 (63.7) |
| <sup>1</sup>Values are n (%) unless otherwise specified.<br><sup>2</sup>Comorbidities were determined from International Classification of Disease billing codes present at the time of the first AF diagnosis, looking back as far as January 1, 2010. Codes were aggregated according to the AHRQ's clinical classification software.<br>AHRQ=Agency for Healthcare Research & Quality; AF=atrial fibrillation; SD=standard deviation |  |

**Figure 1.**
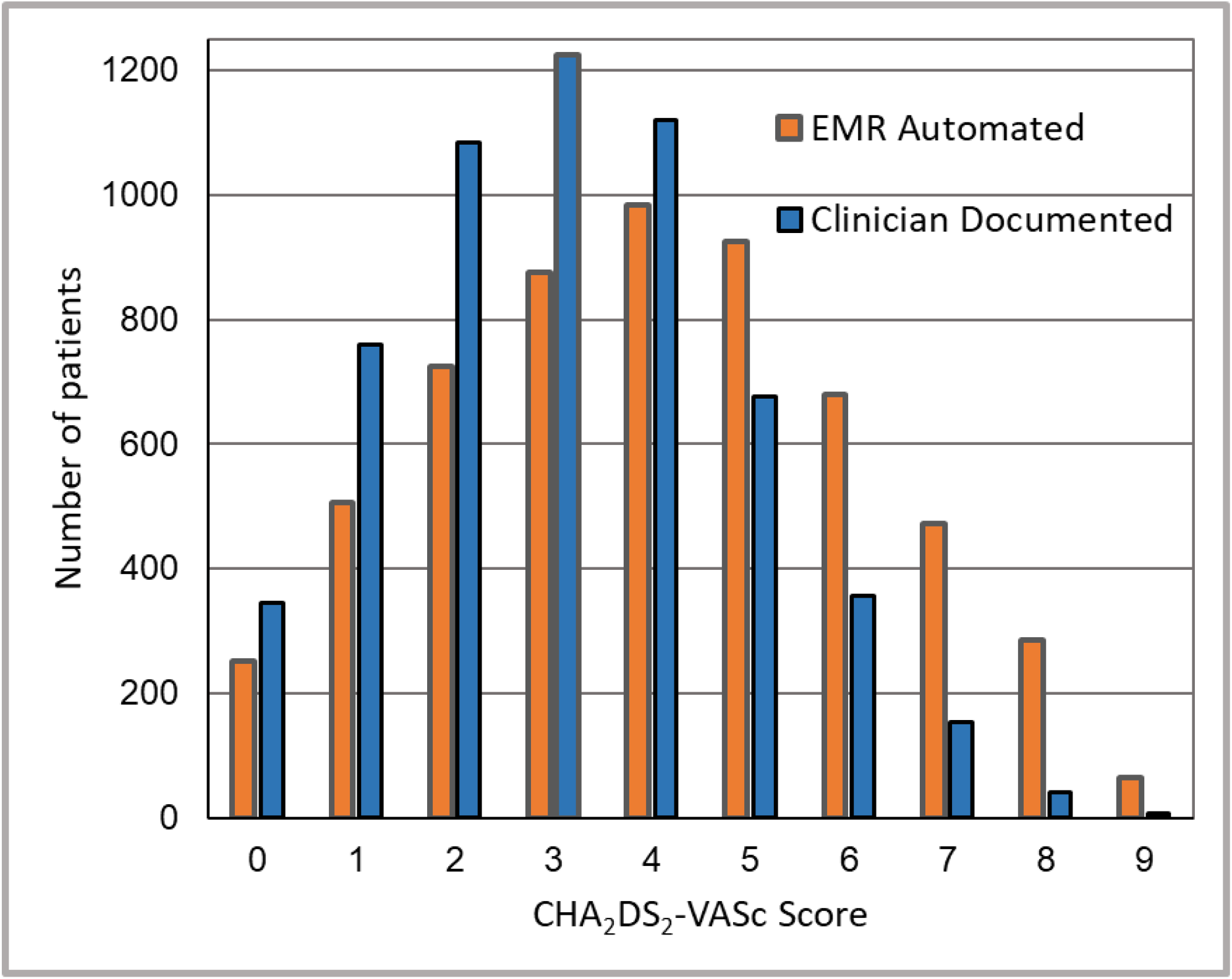
Graphical representations of CHA_2_DS_2_-VASc scores for patients with atrial fibrillation using two methods: EMR automated and clinician documented. The mean CHA_2_DS_2_-VASc score based on the EMR, automated method was 4.05 (SD 2.1, orange bars) compared to 3.13 (SD 1.8, blue bars) based on clinician documentation from the same day (p<0.01). EMR=electronic medical record

The scores differed among the subgroups, and were higher with EMR scores versus documented scores for all subgroups (Table 2). For women, the mean scores were 4.73 (SD 0.04) for the EMR score and 3.68 (SD 0.03) for the documented score (p<0.01 for comparison). For men, the corresponding values were 3.60 (SD 2.1) and 2.76 (SD 1.72; p<0.01). For patients ≤64 years old, the mean EMR score was 2.68 (SD 1.9) compared to 1.89 (SD 1.4) for the documented score (p<0.01). Corresponding values for patients aged 65 to 74 years old were 4.21 (SD 1.9) versus 3.29 (SD 1.5; p<0.01 for comparison). For patients ≥75 years old, the mean EMR score was 5.45 (SD 1.7) compared to the mean documented score of 4.37 (SD 1.4; p<0.01). The mean difference between the two scoring methods was higher for women (1.04 [SD 1.4] for women versus 0.84 [SD 1.27] for men; p<0.01) and younger patients (0.79 [SD 1.2] for age ≤64 years vs 0.91 [SD 1.3] for age 65-74; p<0.002).

**Table 2:** Clinician documented versus EMR automated mean CHA_2_DS_2_-VASc scores among all patients and in selected subgroups.

|  | **Documented Score,**<br>**Mean (SD)** | **Automated score,**<br>**Mean (SD)** | **p-value** |
| --- | --- | --- | --- |
| Overall | 3.13 (1.8) | 4.05 (2.1) | <0.001 |
| Gender |  |  |  |
| Men | 2.8 (1.7) | 3.6 (2.1) | <0.001 |
| Women | 3.7 (1.7) | 4.7 (2.1) | <0.001 |
| Age Categories |  |  |  |
| Age <65 | 1.9 (1.4) | 2.7 (1.9) | <0.001 |
| Age 65-74 | 3.3 (1.5) | 4.2 (1.9) | <0.001 |
| Age 75+ | 4.4 (1.4) | 5.4 (1.7) | <0.001 |

Using a CHA_2_DS_2_-VASc score of ≥2 as the cut-off for OAC treatment, 86.9% (n=5,010) of patients would be treated using the EMR score compared to 80.7% (n=4,663) of patients using the documented scores. If EMR scores were used in lieu of documented scores, 8.3% (n=413) of patients would be re-classified into or out of the treatment group, with 7.2% (n=413) moving up and 1.1% (n=66) of patients moving out of the treatment group (Figures 2 and 3).

**Figure 2:**
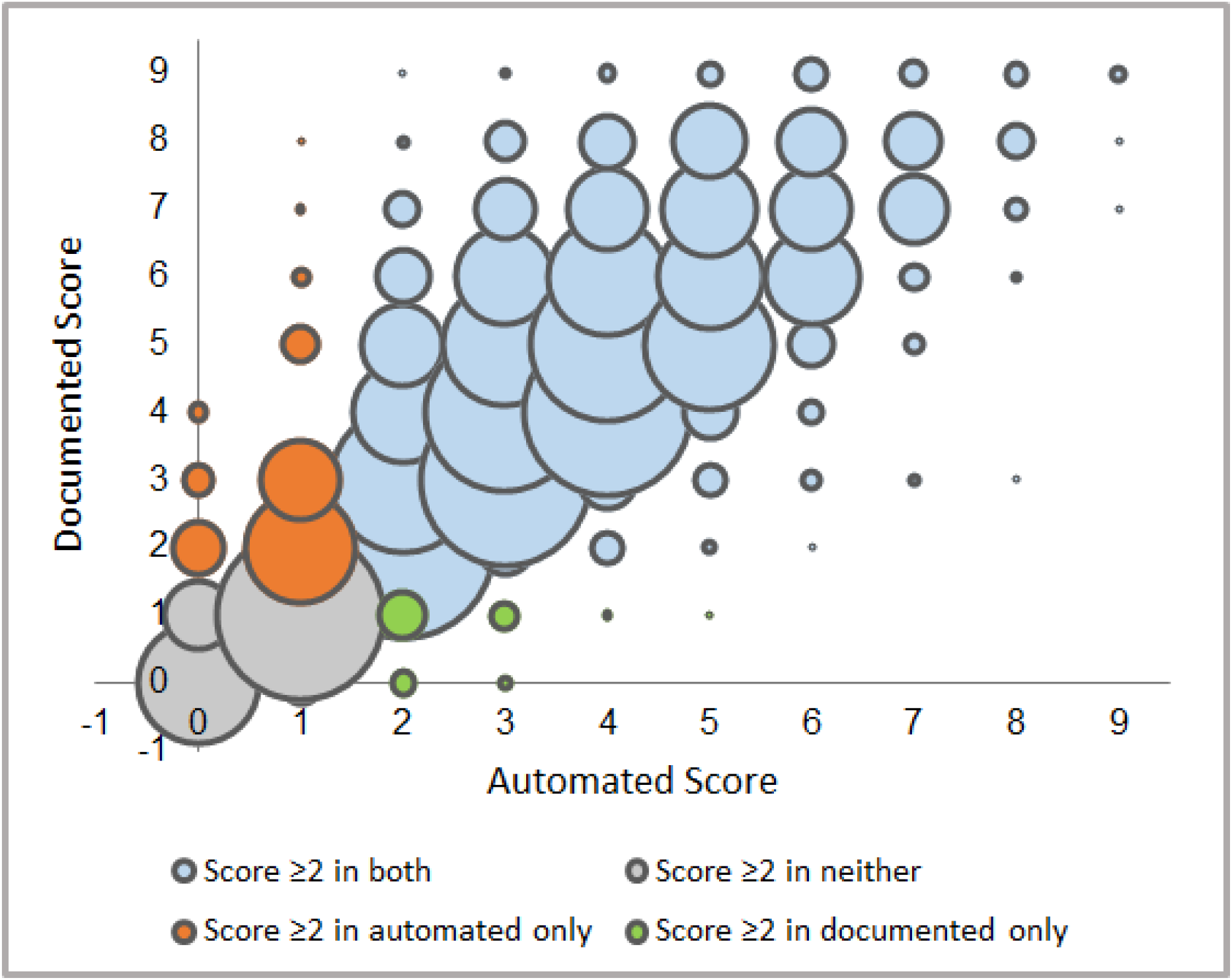
EMR, automated CHA_2_DS_2_-VASc scores plotted against clinician documented CHA_2_DS_2_-VASc scores, with color stratification based a treatment threshold of ≥2. The size of the marker is proportional to the number of patients in that group. This figure plots the EMR, automated CHA2DS2-VASc scores against the clinician document scores, with the size of the circle corresponding to the number of patients in that group. The blue circles indicates agreement, in which the CHA2DS2-VASc scores ≥2 with both methods. The gray circles indicates agreement, in which the CHA2DS2-VASc scores ≥2 with neither method. The orange circles indicates disagreement, in which the CHA2DS2-VASc scores ≥2 with the EMR automated method, but not the clinician documented method (“up classification” with the EMR method). The green circles indicates disagreement, in which the CHA2DS2-VASc scores ≥2 with the clinician documented method, but not the EMR automated method (“down classification” with the EMR method).

**Figure 3:**
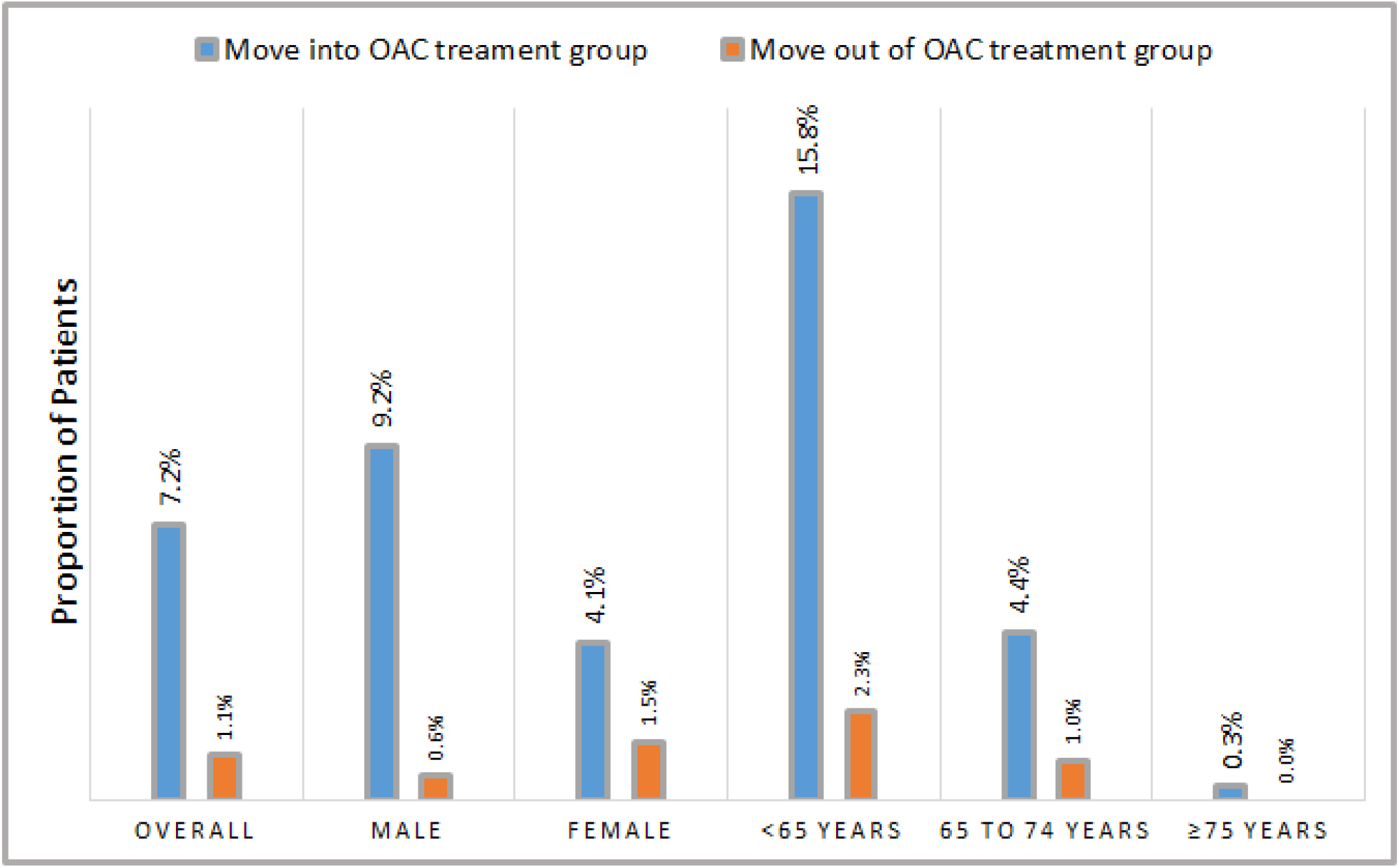
Overall and subgroup re-classification rates if EMR CHA_2_DS_2_-VASc scores were used in lieu of documented scores. If the EMR CHA2DS2-VASc scores were used lieu of clinician documented scores, 7.2% of atrial fibrillation patients would move from a CHA2DS2-VASc of 0 or 1 to a score of ≥2, meaning they would move up into the group for whom oral anticoagulation treatment is recommended. Among the patient subgroups, men and patients ≤64 years old would be most affected, with 9.2% and 15.8% of patients moving up onto the treatment group, respectively. Movement down and out of the treatment group would be infrequent if the EMR scores were used in lieu of the documented scores.

In our subgroup analyses, we found that higher proportions of men and younger patients (≤64 years old) would be re-classified if EMR scores were used in lieu of documented scores. Among 2,322 women, 4.7% (n=109, p<0.01) would be re-classified, with 4.1% (n=95) moving into and 0.7% (n=14) moving out of the treatment group. Among 3,445 men, 10.7% (n=370, p<0.01) would be re-classified, with 9.2% (n=318) moving into and 1.5% (n=52) out of the treatment group. Among 2,060 patients ≤64 years old, 18.1% (n=372, p<0.01) would be re-classified, with 15.8% (n=325) moving into and 2.3% (n=47) moving out of the treatment group. Among 1,877 patients 65-74 years old, 5.4% (n=101, p<0.01) would be re-classified, with 4.4% (n=82) moving into and 1.0% (n=19) moving out of the treatment group. Among 1,830 patients ≥75 years old, <1% would move into to the treatment group and none would move out of the treatment group. (Figure 3).

## Discussion

This study demonstrates that, among AF patients, EMR-based, automated CHA_2_DS_2_- VASc scores matched exactly with clinician-documented scores less than half the time, with the mean EMR score about one point higher than the documented score. When the scores were grouped into low (0-1) and high (≥2) risk, however, agreement between the two methods was over 90%. If the EMR method were used to direct OAC treatment instead of the documented scores, 8.3% of patients would change treatment groups. For the full cohort and sex- and age-based subgroups, the predominant direction was moving up into the OAC treatment group. Specifically, the highest proportion of upward reclassification was for men (almost 1 in 10) and patients ≤64 years old (almost 1 in 5). The proportion of downward reclassification with EMR scores was small, at 1.1% for the whole cohort.

OAC under-treatment in AF is a considerable, widely recognized problem.^2,3,7^ The EMR, with its wealth of data, could streamline CHA_2_DS_2_-VASc score assessment by facilitating automated data aggregation to create points for each component of the score, and thus automatically generate an overall score. On one hand, a well-implemented decision support tool that uses EMR data could help close the treatment gap in AF, although this effect has yet to be proven in limited trials^8,9^. On the other hand, our study raises concerns about EMR-based methods. First, while we reported reclassification rates for the EMR scores compared to documented scores, we do not know which method is correct; the EMR scores could be overly sensitive, capturing “rule-out” diagnoses, or the documented scores could be insensitive because clinicians cannot comprehensively capture the full history. Based on our subgroup analyses, these biases most likely vary between patient groups.

Second, while the scores from the two methods match in some cases, the components of the scores could still differ. For example, the documented score for a 66 year old male could be 2 due to age and hypertension, whereas the EMR score could be 2 due to age and vascular disease. While the importance of component-specific accuracy is uncertain, refinement of EMR-based CHA_2_DS_2_-VASc risk stratification is probably a prerequisite to widespread acceptance and implementation. In these analysis, vascular disease and hypertension were the most prevalent, ICD-based conditions counted in the EMR score, at 56.4% and 76.9%, respectively. While we do not have the specific components that comprised the documented score, these observations suggest that vascular disease and hypertension are good starting points to understand the source of the disparity because false positives in the EMR score may be a concern.

While the EMR and documented score discrepancy was higher for females compared to men, the EMR method would have a greater impact on men: a higher proportion (nearly double the absolute number of men) would move into the OAC treatment group. Similarly, the score discrepancy between the two methods was lower for younger patients, ≤64 years old, but the proportion of patients who would be impacted by EMR based score was higher. These observations imply that an EMR-driven approach would impact treatment among patients without demographic risk factors (older age and female sex) by moving patients into the treatment group. If the goal of a decision support tool is to increase treatment rates, and in light of known “alert fatigue,”^10^ one approach may be to target implementation to these specific subgroups for whom the treatment choice is more likely to change.

Our study is consistent with prior work which suggests that clinician-based stroke risk assessment is lower than risk derived from structured data. A prior report from the Outcomes Registry for Better Informed Treatment of Atrial Fibrillation (ORBIT-AF) Registry found that 72% were classified as high risk when the scores were tallied from the documented conditions, whereas only 16% were deemed high risk by clinician assessment.^11^ An additional issue raised by our work is identifying the “best practice” method to automatically generate CHA_2_DS_2_-VASc scores for AF populations. Older studies using ICD codes suggest >12% misclassification rates,^5^ and ICD coding practices vary between institutions and over time. An alternative may be to extract a value for each component of the score using NLP,^12^ but this approach seems computationally intensive and less practical. Ultimately, a hybrid approach may be best, with a combination of structured data (e.g. ICD codes) and unstructured data (e.g. clinical text).

This study includes several limitations. First, we performed a was a single center analysis; other centers may have less (or more) variation between EMR and documented CHA_2_DS_2_-VASc scores. Second, we relied on clinician documented CHA_2_DS_2_-VASc scores, which biases the population; not all AF patients have a documented CHA_2_DS_2_-VASc target term. Still, we were able to identify over 5,000 patients, which is far greater than could be accomplished with manual chart review to calculate scores. Third, we did not extract each component of the score using NLP, so we could not compare the components of each method to explore the source of discrepancy. Finally, we are reporting theoretical re-classification rates. The actual number of patients who would move up into the treatment group with EMR scores could be smaller due to valid clinical concerns (e.g. bleeding events) or patient preference.

## Conclusions

In conclusion, we found that the EMR-based, automated CHA_2_DS_2_-VASc scores were higher than clinician documented scores, and classified more patients into the OAC treatment group. Ultimately, the painstaking evaluation process described here is required to produce high fidelity EMR-based tools that improve patient outcomes. The discrepancy between scoring methods was particularly notable for men, where almost 1 in 10 patients would be reclassified, and for patients ≤64 years old, where almost 1 in 5 patients would be reclassified. The predominant direction was upward reclassification, such that a substantial proportion of the full AF population would be added to the OAC treatment group; few patients would move out of the treatment group. These results may help tailor targeted interventions for the improvement of population health and research.

## Data Availability

This protected patient information will not be available to others.

## References

1. January CT, Wann LS, Calkins H, Chen LY, Cigarroa JE, Cleveland JC Jr, Ellinor PT, Ezekowitz MD, Field ME, Furie KL, Heidenreich PA, Murray KT, Shea JB, Tracy CM, Yancy CW. 2019 AHA/ACC/HRS Focused Update of the 2014 AHA/ACC/HRS Guideline for the Management of Patients With Atrial Fibrillation. Circulation. 2019;CIR0000000000000665.

2. Shah RU, Rupp AB, Mowery D, Zhang M, Stoddard G, Deshmukh V, Bray BE, Hess R, Rondina MT. Changes in Oral Anticoagulant Treatment Rates in Atrial Fibrillation before and after the Introduction of Direct Oral Anticoagulants. Neuroepidemiology. 2016;47:201– 209.

3. Marzec LN, Wang J, Shah ND, Chan PS, Ting HH, Gosch KL, Hsu JC, Maddox TM. Influence of Direct Oral Anticoagulants on Rates of Oral Anticoagulation for Atrial Fibrillation. J Am Coll Cardiol. 2017;69:2475–2484.

4. Tischer T s., Schneider R, Lauschke J, Nesselmann C, Klemm A, Diedrich D, Kundt G, Bänsch D. Prevalence of atrial fibrillation in patients with high CHADS2- and CHA2DS2VASc-scores: anticoagulate or monitor high-risk patients? Pacing Clin Electrophysiol. 2014;37:1651–1657.

5. Navar-Boggan AM, Rymer JA, Piccini JP, Shatila W, Ring L, Stafford JA, Al-Khatib SM, Peterson ED. Accuracy and validation of an automated electronic algorithm to identify patients with atrial fibrillation at risk for stroke. Am Heart J. 2015;169:39–44.e2.

6. January CT, Wann LS, Alpert JS, Calkins H, Cigarroa JE, Cleveland JC, Conti JB, Ellinor PT, Ezekowitz MD, Field ME, Murray KT, Sacco RL, Stevenson WG, Tchou PJ, Tracy CM, Yancy CW. 2014 AHA/ACC/HRS Guideline for the Management of Patients With Atrial Fibrillation: A Report of the American College of Cardiology/American Heart Association Task Force on Practice Guidelines and the Heart Rhythm Society. Circulation. 2014;130:e199–e267.

7. Hsu JC, Maddox TM, Kennedy K, Katz DF, Marzec LN, Lubitz SA, Gehi AK, Turakhia MP, Marcus GM. Aspirin Instead of Oral Anticoagulant Prescription in Atrial Fibrillation Patients at Risk for Stroke. J Am Coll Cardiol. 2016;67:2913–2923.

8. Arts DL, Abu-Hanna A, Medlock SK, van Weert HCPM. Effectiveness and usage of a decision support system to improve stroke prevention in general practice: A cluster randomized controlled trial. PLoS One. 2017;12:e0170974.

9. Holt TA, Dalton A, Marshall T, Fay M, Qureshi N, Kirkpatrick S, Hislop J, Lasserson D, Kearley K, Mollison J, Yu L-M, Hobbs FDR, Fitzmaurice D. Automated Software System to Promote Anticoagulation and Reduce Stroke Risk: Cluster-Randomized Controlled Trial. Stroke. 2017;48:787–790.

10. Ancker JS, Edwards A, Nosal S, Hauser D, Mauer E, Kaushal R, with the HITEC Investigators. Effects of workload, work complexity, and repeated alerts on alert fatigue in a clinical decision support system. BMC Med Inform Decis Mak. 2017;17:36.

11. Steinberg BA, Kim S, Thomas L, Fonarow GC, Hylek E, Ansell J, Go AS, Chang P, Kowey P, Gersh BJ, Mahaffey KW, Singer DE, Piccini JP, Peterson ED. Lack of Concordance Between Empirical Scores and Physician Assessments of Stroke and Bleeding Risk in Atrial Fibrillation. Circulation [Internet]. 2014 [cited 2019 Sep 16];Available from: https://www.ahajournals.org/doi/abs/10.1161/CIRCULATIONAHA.114.008643

12. Grouin C, Deléger L, Rosier A, Temal L, Dameron O, Van Hille P, Burgun A, Zweigenbaum P. Automatic computation of CHA2DS2-VASc score: information extraction from clinical texts for thromboembolism risk assessment. AMIA Annu Symp Proc. 2011;2011:501–510.

